# The joint effects of exposure to prenatal pesticides and psychosocial factors on epigenetic age acceleration in the first 5 years of life in a South African birth cohort

**DOI:** 10.64898/2026.04.03.26350118

**Authors:** Sarina Abrishamcar, Stephanie M. Eick, Todd Everson, Shakira F. Suglia, M. Daniele Fallin, Robert O. Wright, Syam S. Andra, Jasmin Chovatiya, Ravikumar Jagani, Dana Boyd Barr, Alexandre A. Lussier, Erin C. Dunn, Julie L. MacIsaac, Kristy Dever, Michael S. Kobor, Nadia Hoffman, Nastassja Koen, Heather J. Zar, Dan J. Stein, Anke Hüls

## Abstract

**Background:** Prenatal exposure to pesticides and psychosocial factors often co-occurs, particularly in low- and middle-income settings, yet their joint effects on epigenetic age acceleration (EAA) in early life remain unknown. We investigated the joint associations of prenatal pesticides metabolites and psychosocial factors on EAA in the first five years of life in the South African Drakenstein Child Health Study.

**Methods:** In 643 mothers, we measured 11 urinary pesticide metabolites and seven psychosocial factors during the second trimester of pregnancy. Child DNA methylation was measured in whole blood at ages 1, 3, and 5 years. EAA was estimated using the Horvath, Skin & Blood Horvath (skinHorvath), and Wu epigenetic clocks. Longitudinal associations were estimated using generalized estimating equations, adjusted for confounders. Joint mixture associations were evaluated using weighted quantile sum regression (WQS) and quantile g-computation (QGCOMP).

**Results:** The joint prenatal exposure mixture was positively associated with Wu (β per one quintile increase in the mixture [95% CI]: 0.41 years [0.15, 0.80]), skinHorvath (0.11 years [0.06, 0.16]), and Horvath EAA (0.31 years [0.20, 0.46]) over time using WQS. Psychosocial factors, particularly food insecurity, physical interpersonal violence, and stress biomarkers, contributed most to the total mixture effect for all clocks. Pyrethroid metabolites PBA and TDCCA were top pesticide contributors to Wu EAA. Pathway enrichment analyses of clock-specific CpGs revealed distinct biological architectures, with the Wu clock enriched for neurodevelopmental and immune pathways, and metabolic pathways for the Horvath clock.

**Discussion:** Joint prenatal exposure to pesticides and psychosocial factors was associated with increased EAA across early childhood, with psychosocial factors contributing the most to the total effect. These findings highlight the importance of assessing chemical and non-chemical stressors jointly and clock-specific biological interpretation in epigenetic aging research.

## Introduction

Pregnancy is a critical time window for neurodevelopment, as the developing fetus is uniquely susceptible to environmental toxicants and maternal psychosocial exposures^1,2^. Of particular concern is prenatal exposure to pesticides, many of which are established neurotoxicants and have previously been associated with an increased risk of adverse or delayed developmental outcomes^3–6^. In addition, psychosocial factors, such as maternal depression, psychological distress, socioeconomic distress, and traumatic life events, can dysregulate the maternal-placental-fetal endocrine, immune, and metabolic systems, and have independently been linked to adverse developmental outcomes ^7,8^. Preliminary studies suggest that the harmful effects of environmental toxicants are amplified by psychosocial stressors^8,9^. While psychosocial factors and environmental toxicants have traditionally been studied separately, growing evidence suggests that these exposures may share overlapping biological pathways, including hypothalamic-pituitary-adrenal (HPA) axis dysregulation, oxidative stress, and inflammation. Additionally, exposure to pesticides and psychosocial factors often co-occur, compounded by factors such as increased rates of poverty and structural inequalities which disproportionately burden communities of low socioeconomic status and especially those in low- and middle-income countries (LMICs)^10,11^.

Epigenetic alterations are one biological mechanism hypothesized to explain how the combined exposure to environmental and psychosocial factors impact child health outcomes^12–16^. Epigenetic age acceleration (EAA) is a well-established biomarker of biological aging which assesses differences between estimated epigenetic age and actual chronological age^17^. Recent studies have suggested that epigenetic age acceleration in pediatric populations may be associated with exposure to environmental and psychosocial factors, as well as developmental milestones^15,16,18–20^. However, to date, the combined impact of pesticide exposure and psychosocial factors on epigenetic age acceleration across early childhood has not been studied. In adults, increased EAA is generally associated with elevated risk of morbidity and mortality^17^. By contrast, in childhood, the direction and significance of EAA, and whether it is adaptive or maladaptive at certain periods of development, remain unclear^21,22^. Because early childhood represents periods of rapid biological change and development, repeated measures of EAA may provide insight into early-life developmental processes that cannot be captured at a single timepoint. Most existing studies have assessed EAA at a single timepoint, and have been conducted primarily in high income populations, and no study has examined joint exposure to both pesticides and psychosocial factors in relation to EAA in LMIC settings.

Here, we investigated the joint effects of prenatal pesticide urinary metabolites and psychosocial factors on childhood EAA measured at one, three, and five years of age in the Drakenstein Child Health Study, a well-characterized South African birth cohort. We focus on the Horvath^23^, Wu^24^, and Skin and Blood Horvath (skinHorvath)^25^, since these three epigenetic clocks incorporated peripheral blood samples from childhood in their training data, with the Wu clock specifically created for childhood EAA, and the two Horvath clocks created as life course EAA clocks. We utilized two complementary mixture methods, weighted quantile sum regression (WQS) and quantile-based g-Computation (QG-COMP) to characterize the joint effects of prenatal exposures on EAA across early childhood, and to identify mixture components that contribute most to the total effect. We also conducted pathway enrichment analysis on each epigenetic clock’s set of CpG sites to investigate the underlying biological processes represented by each measure of epigenetic age and whether clock-specific biological architecture may partly explain the distinct associations with environmental exposures.

## Methods

### Study population

The Drakenstein Child Health Study (DCHS) is a well-characterized, population-based birth cohort study based in a peri-urban region outside Cape Town, South Africa. The cohort, which has previously been described in detail, is comprised of mother-child pairs of Black African and mixed race ^26,27^. Consenting pregnant women (N=1137) were recruited during their second trimester of pregnancy from two primary health clinics, TC Newman and Mbekweni, between 2012 and 2015. Mother-child pairs have been followed closely from birth up to at least 10 years. Ethical approval was given by the Human Research Ethics Committee of the Faculty of Health Sciences at the University of Cape Town. Written consent for participation was obtained from the mother on behalf of herself and the child and mothers were reconsented annually.

### DNA methylation measurements

Whole blood was extracted using the Qiagen DNeasy Blood & Tissue kit. DNA methylation (DNAm) was measured from whole blood using the Infinium MethylationEPIC v2.0 BeadChip assay at age one (n=652), three (n=708), and five (n=760) years. Samples were randomized across chips and plates to remove potential confounding by plate, chip, or row, while maintaining samples from the same participant on the same chip. DNAm preprocessing and quality control were completed using the *meffil* R package, as previously described^28^. Probes with a bead count of less than three (n=19) or a detection p-value <0.1 (n=1,475) were removed. Samples were considered outliers and removed if their predicted mean methylation was more than three standard deviations from the expected methylation values (19 samples). Sex mis-matched samples were also removed (8 samples). A total of 1,494 probes and 28 samples were removed, leaving 929,106 probes and 2092 samples for downstream analysis. The data were functionally normalized. The first 15 principal components were calculated from the raw and normalized DNAm data to identify batch effects, and none were detected.

Cell-type proportions were estimated using the UniLife reference panel from the *EpiDISH* R package, optimized to predict blood-based immune-cell fractions for any age group and specifically for longitudinal studies that include infants and children^29,30^. UniLife predicts 19 immune cell-types including seven cord-blood subtypes (B cells, NK cells, granulocytes, monocytes, nRBCs, CD4T, and CD8T cells) and 12 adult subtypes (naïve and mature B cells, naïve and mature CD4T cells, naïve and mature CD8T cells, T-regulatory cells, NK cells, neutrophiles, monocytes, eosinophils, basophils).

EAA was calculated as the residuals from regressing epigenetic age (EA) on chronological age. A positive value indicates increased age acceleration, in which a person’s EA is predicted to be older than their chronological age. We calculated EA estimates using the following epigenetic clock algorithms that have been trained using datasets including children and have been validated for use in pediatric populations^17^: Wu’s clock^24^, Horvath’s pan-tissue clock^23^, and Horvath’s Skin and Blood clock^25^. All three clocks were calculated using the *methylclock* Github release^31^.

### Urinary pesticide metabolites

Pesticide metabolites were measured in biobanked urine samples collected from mothers during the second trimester of pregnancy. Urine samples were collected using a spot, clean catch method into sterile polypropylene collection tubes. Samples were transported, processed, aliquoted, and stored frozen in the DCHS research laboratory and curated in the DCHS biorepository using the Freezerworks inventory system. The biomarker measurements were provided by the Human Health Exposure Analysis Resource (HHEAR) Targeted Analysis Laboratory at the Icahn School of Medicine at Mount Sinai in New York, NY, using a previously developed and validated bioanalytical method^32^. A multiclass liquid chromatography tandem mass spectrometry (LC-MS/MS) analytical method was used which included the quantification of 7 organophosphate insecticide metabolites, one neonicotinoid insecticide metabolite, two pyrethroid insecticide metabolites, and two chlorinated phenols. Quality control consisted of experimental blanks (reagent- and matrix-based), matrix spikes at low, medium, and high validation levels, investigator-initiated paired duplicate samples blinded to the lab, two HHEAR QC urine pools (three of each analyzed per 100 samples), NIST standard reference materials 3672 and 3673, and archived, previously characterized proficiency-testing material, as outlined in HHEAR^33^.

### Psychosocial factors

Measures of psychosocial factors were obtained from mothers during the second trimester of pregnancy, including depressive symptoms, psychological distress, traumatic life events, substance use, socioeconomic status, perceived household food insecurity, and interpersonal violence. We used continuous, total scores for each psychosocial factor by summing the items within each scale, with higher values indicating greater exposure or severity.

Depressive symptoms were measured using the Edinburgh Postnatal Depression (EPDS) scale, which is designed to measure postnatal depression^34^ and has also been verified for prenatal depression in an African setting^35,36^. It consists of 10 items with scores ranging from 0 to 30. Psychological distress was measured using the WHO-endorsed Self-Reporting Questionnaire (SRQ-20) which consists of 20 items with scores ranging from 0 to 20^35^. Traumatic life events were assessed using the World Mental Health Life Events Questionnaire, used to assess exposure to 13 areas of stressful or negative life events^37,38^. This scale consists of 17 items with scores ranging from 0 to 17.

Socioeconomic status (SES) was defined as a summed composite score of multiple SES variables: maternal employment status, maternal education, household income, and household assets, as previously used and validated in this cohort^39^. Perceived household food insecurity was measured using an adapted version of the USDA Household Food Security Scale ^40, 41,42^. This measure has been previously validated in this cohort^40^.

Prenatal substance use was measured using the WHO-derived Alcohol, Smoking, and Substance Involvement Screening Test (ASSIST)^43^. The ASSIST questionnaire consists of 8 items assessing alcohol and substance use with scores ranging from 0 to greater than 26. Individual items were summed to obtain a separate continuous score for alcohol and tobacco use, with higher scores indicating greater risk for substance-related health problems. The ASSIST questionnaire has been validated for use in South Africa^43^. Interpersonal violence was measured using the Intimate Partner Violence (IPV) questionnaire ^44,45^ which assesses lifetime and recent exposure to physical and emotional interpersonal violence. A separate continuous score was obtained for emotional IPV and physical IPV by summing all items in each domain.

In addition, biomarkers of psychosocial stress were measured using prenatal urinary cortisol and cortisone, which were quantified together with pesticide metabolites in the same urine aliquot and validated multiclass assay, following HHEAR standards as described above^32,33^. We additionally calculated the cortisol-to-cortisone ratio as a marker of 11β-hydroxysteroid dehydrogenase type 2 (11β -HSD2) activity, the enzyme that converts active cortisol to inactive cortisone. Higher ratios indicate more circulation of active cortisol and have been used as indicators of stress-related HPA axis dysregulation^46^.

### Statistical Analysis

All analyses were restricted to participants with complete prenatal urinary pesticide metabolite measurements (n=643). Urinary pesticide metabolites that were greater than the limit of detection (LOD) in at least 33% of study samples were included in the analysis, resulting in 11 pesticide metabolites. These were retained as continuous variables in mixture models, where the joint distribution of exposures is modeled simultaneously and sparsity in any single component has less influence on the overall mixture estimate. Concentrations below the LOD were treated as left-censored and imputed using the *mice.impute.leftcenslognorm* function from the *qgcomp* R package, which draws from a left-censored log-normal distribution^47^.

Non-normal variables were log-transformed on the natural log scale, and a constant of 1 was added to psychosocial survey scales that had zero-values. The SES scale was reversed so that a higher score indicated lower socioeconomic status, in line with all other exposures in which a higher score indicates greater exposure or severity.

All models were adjusted for mother’s self-reported race (Black African/Mixed), maternal age at enrollment, and maternal HIV status (Infected/Not infected), as defined as the minimally sufficient set of confounders from a directed acyclic graph (DAG). Child age and child sex were included as precision covariates and prenatal urinary creatinine concentration was included as a covariate in models that included pesticide metabolites or psychosocial stress biomarkers to account for variation in urine dilution^48^.

#### Multiple imputation of missing values

Missing data were assumed to be missing at random (MAR), so we used multiple imputation by chained equations (MICE) using the *mice* R package to impute missing values in covariates and EA measures^49^. Five imputed datasets with 50 iterations were generated using predictive mean matching for continuous variables, left-censored log-normal for pesticide values below LOD, and polytomous regression for categorical variables. Because of the complexity of downstream mixture analyses that do not readily support pooling via Rubin’s rules, one imputed dataset was selected at random for primary analyses. To evaluate the stability of the imputed datasets, we repeated the single exposure models in the remaining four imputed datasets as a sensitivity analysis.

#### Single exposure models

We conducted single exposure longitudinal models using generalized estimating equations (GEE), a semi-parametric approach that estimates the population-average associations using a marginal model. We specified an exchangeable working correlation structure to account for within-child clustering of repeated measures. Standard errors were computed using the empirical (sandwich) variance estimator.

#### Mixture models

We utilized two environmental mixture methods to assess the joint effects of prenatal pesticide and psychosocial exposure on epigenetic age acceleration over time, including weighted quantile sum regression (WQS)^50^ and quantile-based g-Computation (QG-COMP)^47^. All 11 prenatal urinary pesticide metabolites and seven psychosocial factors were included in the exposure mixture.

WQS estimates the effect of a one quintile increase in the exposure mixture on EAA over time. WQS regression works by calculating an empirically weighted index, in which exposures with weaker effects on the outcome have a smaller weight and exposures with stronger effects on the outcome have a larger weight. We incorporated repeated holdout validation to randomly partition the dataset 100 times to improve result stability in our smaller sample size^51^. The longitudinal extension of WQS fits linear mixed-effects models in each of the 100 repeated holdout datasets to accommodate intra-subject correlated outcomes^52^. We accounted for within-child correlation by including a subject-specific random intercept for each participant. The summary statistics were pooled across all datasets, with the final effect estimate defined as the 50^th^ percentile and the 95% confidence intervals defined as the 2.5^th^ and 97.5^th^ percentiles.

Complementary to WQS, QG-COMP estimates the effect of simultaneously increasing all exposures in the mixture by one quintile on EAA over time using GEE. Unlike WQS, which has a directional homogeneity assumption such that all exposures are acting in the same direction, QG-COMP allows for exposures to have varying directions of effect. Applying both approaches allows for validation of findings across methods; if both approaches identify similar key exposures acting in the same direction, it suggests that the WQS directional homogeneity assumption is not violated.

In secondary analyses, we constructed WQS and QG-COMP models individually for each of the domains, a pesticide metabolite mixture and a psychosocial factor mixture, to assess whether one exposure domain was contributing more to the total effect.

#### Sensitivity analyses

We conducted several sensitivity analyses to evaluate the robustness of our findings. For the single-exposure longitudinal models, we (1) additionally adjusted for estimated immune blood cell-type proportions to assess how much of the total effect was driven by cell compositions, (2) assessed the accumulating effect that each exposure has on the pace of EAA over time by including an interaction term between each exposure and time, represented by chronological age, (3) assessed whether the association between each exposure and EAA differed by child sex by including an interaction term between each exposure and sex, (4) assessed the robustness of our findings by dropping metabolites that were detected in less than 50% of samples, and (5) repeated the main analyses across the four additional imputed datasets.

### Pathway enrichment analysis of epigenetic clocks

The Horvath, Wu, and Skin and Blood Horvath epigenetic clock algorithms were trained on distinct sets of CpG sites and therefore may capture different underlying biological processes. To investigate the biological processes represented by each EA clock, we conducted Gene Ontology (GO) enrichment analysis separately for the CpG sites comprising each clock using the *gometh* function from the *missMethyl* R package^53^. This included 340/353 available CpGs from Horvath, 107/111 CpGs from Wu, and 374/391 CpGs from skinHorvath. The background set for enrichment testing was restricted to CpGs present on both Illumina 27K and 450K arrays for the Horvath and Wu clocks, and present on both the 450K and EPICV1 array for the skinHorvath clock, consistent with the platforms used in training. As our goal here was not statistical inference, but rather to identify broader themes of biological processes, we restricted inference to Biological Process (BP) GO terms and filtered the resulting GO terms by a raw p-value < 0.05 and at least two annotated genes per term. To facilitate interpretation and comparison across clocks, we grouped GO terms into broader functional categories based on shared keywords and biological themes using a large language model (LLM; ChatGPT 5.0) using a structured prompt. The assigned categories were manually reviewed to confirm accuracy. This approach enabled us to characterize both overlapping and clock-specific biological processes underlying each EA clock.

## Results

### Study population characteristics

This analytic sample included 643 mother-child pairs with complete prenatal pesticide metabolite measurements and methylation data from children at 12 months (n=448), 36 months (n=511), and 60 months (n=583) (Table 1). Study characteristics were evenly distributed across timepoints. Approximately 47% of children were female and the cohort was evenly split between Black African (50%) and Mixed race (50%). The mean maternal age at enrollment was 26.8 years, the mean gestational age at delivery was 38.6 weeks, and approximately 22% of mothers were living with HIV. In terms of socioeconomic indicators, approximately one third of mothers had completed secondary education, over 85% of households earned less than R5000 per month, and nearly half of mothers were unemployed. Psychosocial factors were evenly distributed across timepoints, with low levels of missing data (<7% for all measures).

**Table 1:**
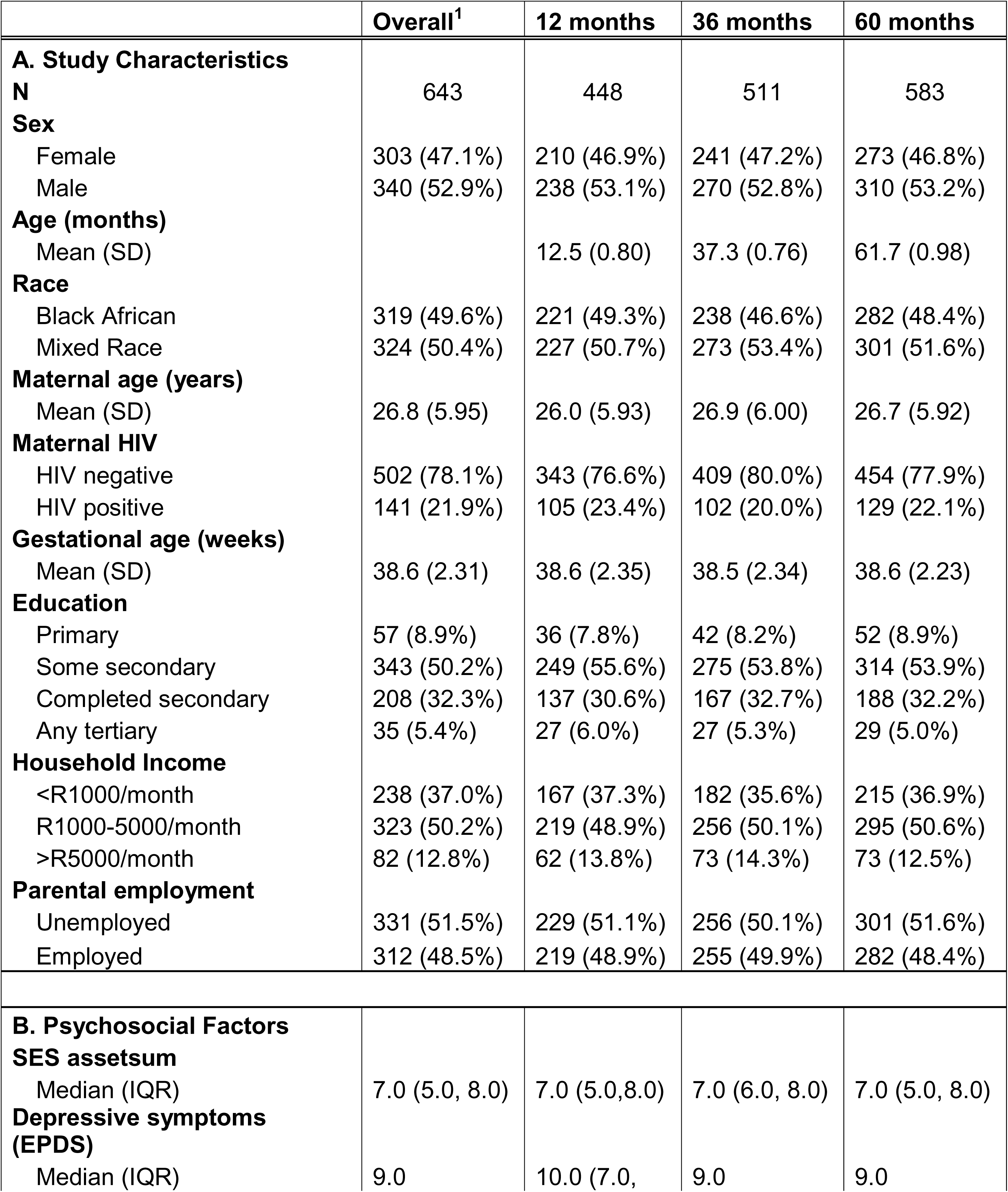
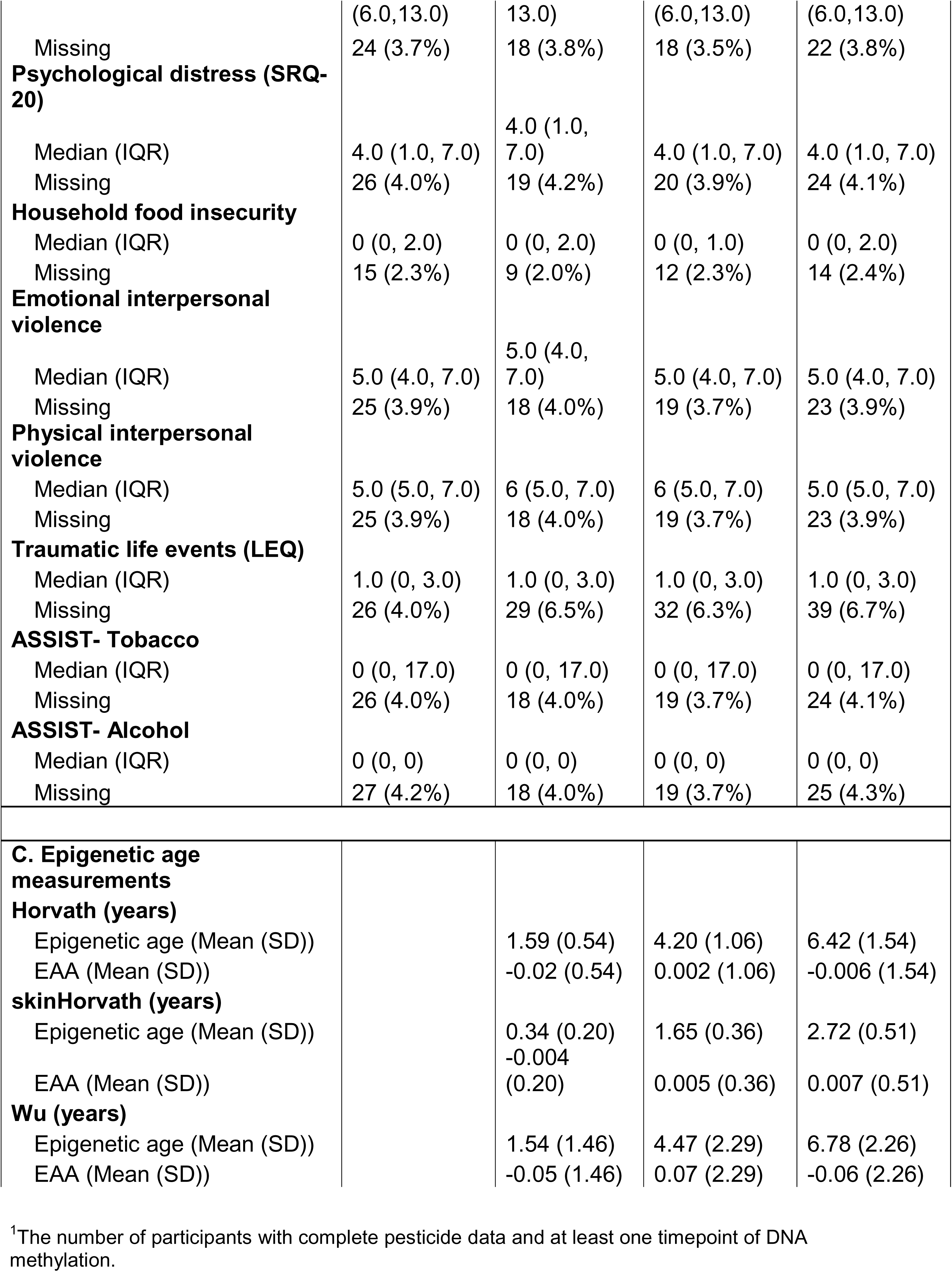
Population characteristics, psychosocial factors, and epigenetic age measurements at 12, 36, and 60 months. Drakenstein Child Health Study, Cape Town, South Africa, 2012-2015.

The proportion of adult immune cell-types increased, while the proportion of cord-blood immune cell-types decreased across the three time points. At age 12 months, roughly half of the cell-type proportions are cord-blood subtypes and half are adult subtypes. By age 60 months, approximately 75% of the cell-type proportions are estimated to be adult immune cell-types (Figure S1).

The correlation between epigenetic age and chronological age varied across clocks (Figure S2). The Horvath clocks had the highest prediction accuracy (Horvath R^2^=0.74, skinHorvath R^2^=0.86) whereas the Wu clock had the lowest (R^2^=0.51). Pesticide metabolite concentrations were all positively correlated with each other, with the highest Pearson correlation being between two pyrethroid metabolites, trans-3-(2,2-dichlorovinyl)-2,2-dimethylcyclopropane-l-carboxylic acid (TDCCA) and 3-phenoxybenzoic acid (PBA) (r=0.62) (Figure S3). Nine of the eleven pesticide metabolites were detected in ≥50% of the participants, with 6-chloronicotinic acid (CINA6) and 2,4-dichlorophenol (D24) being detected in 35% and 45% of participants, respectively. The stress biomarkers cortisone and cortisol were detected in ≥ 99% of participants (Table 2). Psychosocial factors were correlated multi-directionally with each other (Figure S4).

**Table 2:** Distribution of prenatal urinary pesticide metabolites and stress biomarkers measured at the second trimester of pregnancy, Drakenstein Child Health Study, Cape Town, South Africa, 2012-2015. Shown are the percentage of samples above the limit of detection (% ≥ LOD), geometric mean (GM) and geometric standard deviation (GSD), and the chemical concentrations at the 25^th^ percentile, median, and 75^th^ percentile. Values below the LOD are indicated as <LOD. Concentrations are reported in ng/ml.

| Metabolite | % ≥ LOD | GM (GSD) | Median [IQR] | | |
| --- | --- | --- | --- | --- | --- |
|  |  |  | 25<sup>th</sup> | Median | 75<sup>th</sup> |
| Organophosphates |  |  |  |  |  |
| diethyl phosphate (DEP) | 94% | 2.61 (3.00) | 1.38 | 2.67 | 4.78 |
| diethyl thiophosphate (DETP) | 82% | 0.96 (2.95) | 0.44 | 0.90 | 1.64 |
| dimethyl dithiophosphate (DMDP) | 54% | 0.45 (3.24) | <LOD | 0.46 | 0.93 |
| dimethyl phosphate (DMP) | 98% | 10.85 (3.42) | 4.74 | 11.70 | 23.45 |
| dimethyl thiophosphate (DMTP) | 99% | 6.08 (2.29) | 3.31 | 5.49 | 9.99 |
| p-nitrophenol (PNP) | 73% | 0.95 (2.65) | <LOD | 0.89 | 1.65 |
| Pyrethroids |  |  |  |  |  |
| 3-phenoxybenzoic acid (PBA) | 60% | 0.53 (2.92) | <LOD | 0.55 | 1.08 |
| trans-3-(2,2-dichlorovinyl)-2,2-dimethylcyclopropane-l-carboxylic acid (TDCCA) | 53% | 0.44 (3.06) | <LOD | 0.34 | 0.83 |
| Neonicotinoid |  |  |  |  |  |
| 6-chloronicotinic acid (CINA-6) | 35% | 0.22 (3.23) | <LOD | <LOD | 0.43 |
| Organochlorines |  |  |  |  |  |
| 2,4-dichlorophenol (D24) | 45% | 0.19 (3.02) | <LOD | <LOD | 0.37 |
| 2,4,5-trichlorophenol (TCP) | 95% | 2.57 (2.74) | 1.31 | 2.66 | 5.04 |
| Stress Biomarkers |  |  |  |  |  |
| Cortisone | 100% | 250.07 (1.75) | 183.50 | 257.00 | 346.00 |
| Cortisol | 99% | 80.88 (2.93) | 45.00 | 94.00 | 170.50 |
| Cortisol:Cortisone<sup>1</sup> | -- | 0.33 (2.51) | 0.21 | 0.37 | 0.59 |

LOD=limit of detection; GM=geometric mean; GSD=geometric standard deviation
<sup>1</sup>The cortisol:cortisone ratio is a derived variable calculated from measured cortisol and cortisone concentrations. %>LOD is not applicable, and values are unitless.

### Single exposure models

We assessed single-exposure associations between each prenatal pesticide metabolite concentration or psychosocial factor and EAA during follow-up using adjusted GEE models (Figure 1A; Tables S1-S3) and at 1, 3, and 5 years individually using adjusted linear regression models (Figure 1B; Tables S4-S6). All effect estimates are presented as the change in EAA per one interquartile range (IQR) increase in the natural log-transformed exposure. Three psychosocial factors were associated with Horvath EAA during follow-up. Traumatic life events (β=0.26 years, 95% CI: 0.12, 0.39), cortisol (β=0.11 years, 95% CI: 0.02, 0.19), and cortisone (β=0.08 years, 95% CI: 0.004, 0.15) were associated with increased Horvath EAA. One psychosocial factor and two pesticide metabolite concentrations were associated with Wu EAA during follow-up. Physical IPV, PBA (β=0.30 years, 95% CI: 0.06, 0.53) and TDCCA (β=0.23 years, 95% CI: 0.04, 0.42) were associated with increased Wu EAA. Three psychosocial factors were associated with skinHorvath EAA during follow-up. Food insecurity (β=0.05 years, 95% CI: 4.58E-03, 0.09), cortisol (β=0.05 years, 95% CI: 0.02, 0.08), and cortisol:cortisone (β=0.04 years, 95% CI: 0.01, 0.06) were associated with increased skinHorvath EAA. The magnitudes and directions of effect were similar across clocks for the psychosocial factors, with most psychosocial factors being positively associated with EAA. Differences were more apparent for pesticides, with pesticides being generally positively associated with the Wu clock and not associated with Horvath and skinHorvath clocks.

**Figure 1:**
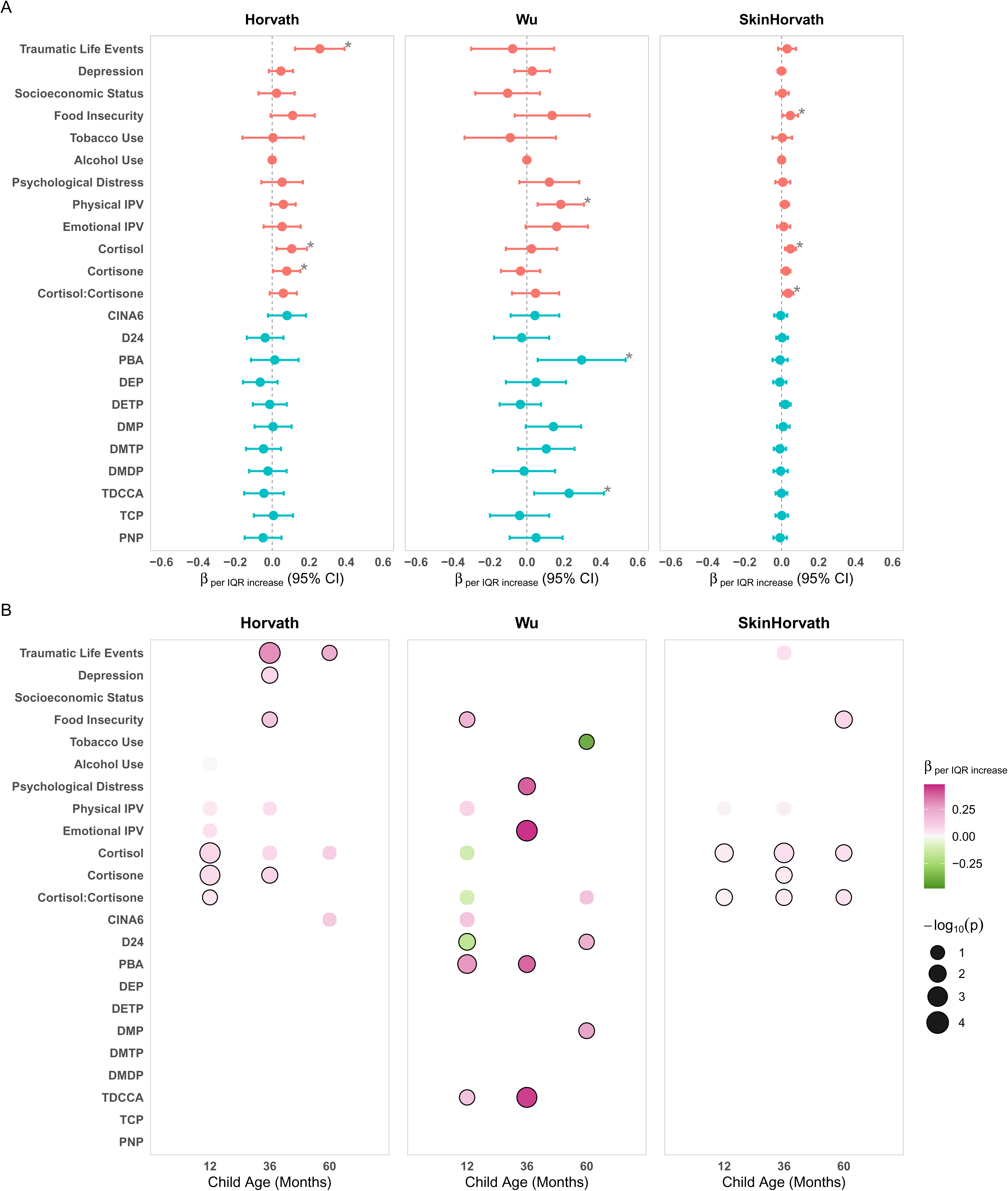
Single-exposure associations between prenatal pesticide metabolites and psychosocial factors and epigenetic age acceleration (Horvath, Wu, and Skin and Blood Horvath EAA) across the first five years of life. Row. **A.** Longitudinal associations between each prenatal exposure and EAA across 12, 36, and 60 months estimated using GEE models. Exposures are log transformed Psychosocial factors are shown in salmon, and pesticide metabolites are shown in blue. Asterisks denote significant associations at p < 0.05. **Row B.** Age-specific associations between prenatal exposures and EAA estimated individually at 12, 36, and 60 months using linear regression models. Circles are shown only for associations with p < 0.10. Circle color represents the direction and magnitude of the beta coefficient per IQR increase in log-transformed exposure, with pink indicating positive associations and green indicating negative associations. Circle size corresponds to –log_₁₀_(p-value), such that larger circles indicate stronger statistical evidence. Circles outlined in black denote associations with p < 0.05. All models were adjusted for child age, child sex, race, maternal age at enrollment, and maternal HIV status. Models including pesticide metabolites or psychosocial biomarkers were additionally adjusted for urinary creatinine.

Associations varied across childhood developmental timepoints and differed by epigenetic clock type.. While patterns varied by age, the greatest number of associations were identified in the first 3 years of life and tapered off at age 5 years. For example, physical IPV and cortisone were associated with increased Horvath EAA at 12 months (physical IPV: β=0.04 years, 95% CI: −0.003, 0.08; cortisone: β=0.08 years, 95% CI: 0.03, 0.13) and 36 months (physical IPV: β=0.07 years, 95% CI: −0.01, 0.15; cortisone: β=0.11 years, 95% CI: 0.02, 0.20), but not at 60 months (physical IPV: β=0.07, 95% CI: −0.04, 0.19; cortisone: β=-0.06 years, 95% CI: , 0.20). Additionally, PBA and TDCCA were associated with increased Wu EAA at 12 months (PBA: β=0.30, 95% CI: 0.11, 0.50; TDCCA: β=0.18, 95% CI: 0.01, 0.34) and 36 months(PBA: β=0.40, 95% CI: 0.09, 0.72; TDCCA: β=0.47, 95% CI: 0.20, 0.73), but not at 60 months (PBA: β=0.19, 95% CI: −0.12, 0.49; TDCCA: β=0.05, 95% CI: −0.21, 0.31).

Overall, pesticides were associated increased Wu EAA and psychosocial factors were largely associated with increased EAA across all three clocks.

### Joint mixture models of pesticides and psychosocial factors

The WQS index was positively associated with Horvath EAA (β=0.31 years per one quintile increase in the exposure mixture, 95% CI: 0.20, 0.46), Wu EAA (β=0.41 years, 95% CI: 0.15, 0.80), and skinHorvath EAA (β=0.11 years, 95% CI: 0.06, 0.16) during follow-up (Figure 2A). Overall, psychosocial factors accounted for the largest proportion of the total mixture weight. Across all three clocks, food insecurity, physical IPV, tobacco use, and biomarkers of stress were top contributors to the total mixture effect. For Horvath EAA, the top five contributors to the total effect were food insecurity, traumatic life events, cortisol, tobacco use, and physical IPV (Figure 2B). For Wu EAA, food insecurity, physical IPV, PBA, alcohol and tobacco use contributed most to the total effect. For skinHorvath EAA, the top five contributors were food insecurity, cortisol:cortisone, cortisone, physical IPV, and tobacco use. Although pesticide metabolites generally had smaller weights, they contributed more prominently to Wu EAA compared with Horvath and skinHorvath EAA. For Wu EAA, PBA, DMP, and TDCCA were among the ten largest weights. By contrast, CINA6 was the only pesticide metabolite that substantially contributed to Horvath EAA, whereas diethyl thiophosphate (DETP) was the primary pesticide contributor to skinHorvath EAA.

**Figure 2:**
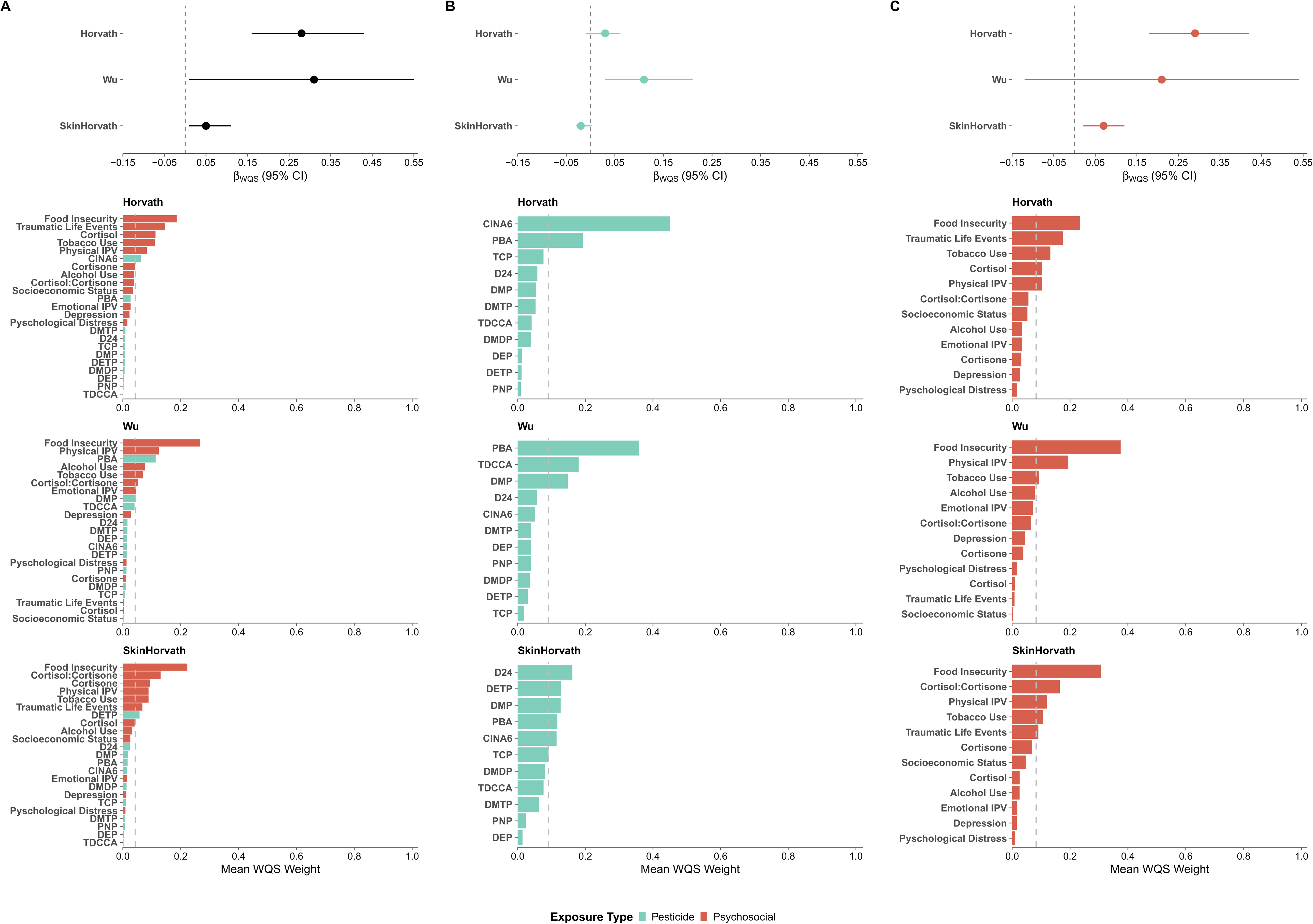
Weighted quantile sum (WQS) mixture associations between prenatal pesticide metabolites and psychosocial factors and epigenetic age acceleration (Horvath, Wu, and Skin and Blood Horvath) during follow-up. Column. **A.** Association between the joint prenatal exposure mixture and EAA across 12, 36, and 60 months. **Column B.** Longitudinal associations between pesticide metabolite mixtures alone and EAA. **Column C.** Longitudinal associations between psychosocial factor mixtures alone and EAA. Results are shown from the longitudinal mixed-effects WQS regression with repeated holdout validation. Beta estimates represent the change in EAA (years) per one quintile increase in the exposure. Point estimates denote pooled effect estimates (median across 100 repeated holdouts) and the corresponding 95% confidence intervals (2.5th–97.5th percentiles). For each column, lower plots display the corresponding WQS mixture weights indicating the relative contribution of each exposure to the overall mixture effect. Weights sum to one within each model. Pesticide metabolites are shown in blue and psychosocial factors are shown in salmon. All models were adjusted for child age, child sex, race, maternal age at enrollment, and maternal HIV status. Models including pesticide metabolites or psychosocial biomarkers were additionally adjusted for urinary creatinine.

Using QGCOMP, the overall mixture, while consistently in the positive direction, was not associated with EAA for any of the clocks (Horvath: Ψ=0.24 years per one quintile increase in the exposure mixture, 95% CI: −0.10, 0.59, Wu: Ψ=0.57 years, 95% CI: 0.01, 1.14, skinHorvath Ψ=0.12 years, 95% CI: −0.02, 0.25) (Figure S5A). The top contributors were consistent with WQS.

### Domain-specific mixture models

For WQS and QGCOMP mixture models, we conducted stratified analyses for the pesticides and psychosocial factors to assess whether one exposure domain was contributing more to the total effect. The pesticide mixture was associated with increased Wu EAA, whereas the psychosocial factors mixture was associated with increased Horvath and skinHorvath EAA.

With WQS, there was an association between the pesticide mixture and increased Wu EAA (β=0.16 years, 95% CI: 0.03, 0.27) during follow-up (Figure 2A). No association was detected with Horvath EAA (β=0.01 years, 95% CI:-0.06, 0.07) or skinHorvath EAA (β= −0.01 years, 95% CI: −0.03, 0.01). The top contributor for the Horvath clock was CINA6 and PBA, the top contributors for the Wu clock were PBA, TDCCA, and DMP, and the top contributors for the skinHorvath clock were D24, DETP, and DMP. These are consistent with the top pesticide contributors in the joint mixture model.

For psychosocial factors, the mixture was associated with increased Horvath (β= 0.31 years, 95% CI: 0.20, 0.45) and skinHorvath EAA (β= 0.07 years, 95% CI: 0.02, 0.12) during follow-up. (Figure 3B). The association for Wu EAA was not significant, however the effect estimate was also in the positive direction (β= 0.33 years, 95% CI: −0.02, 0.61). The top contributors for Horvath EAA were food insecurity, traumatic life events, tobacco use, cortisol, and physical IPV. The top contributors for skinHorvath were food insecurity, cortisol:cortisone, physical IPV, and tobacco use. The top contributors for Wu were food insecurity, physical IPV, and tobacco use. Similar to the joint mixture model, food insecurity, physical IPV, tobacco use, and biomarkers of stress were the largest contributors across clocks.

**Figure 3:**
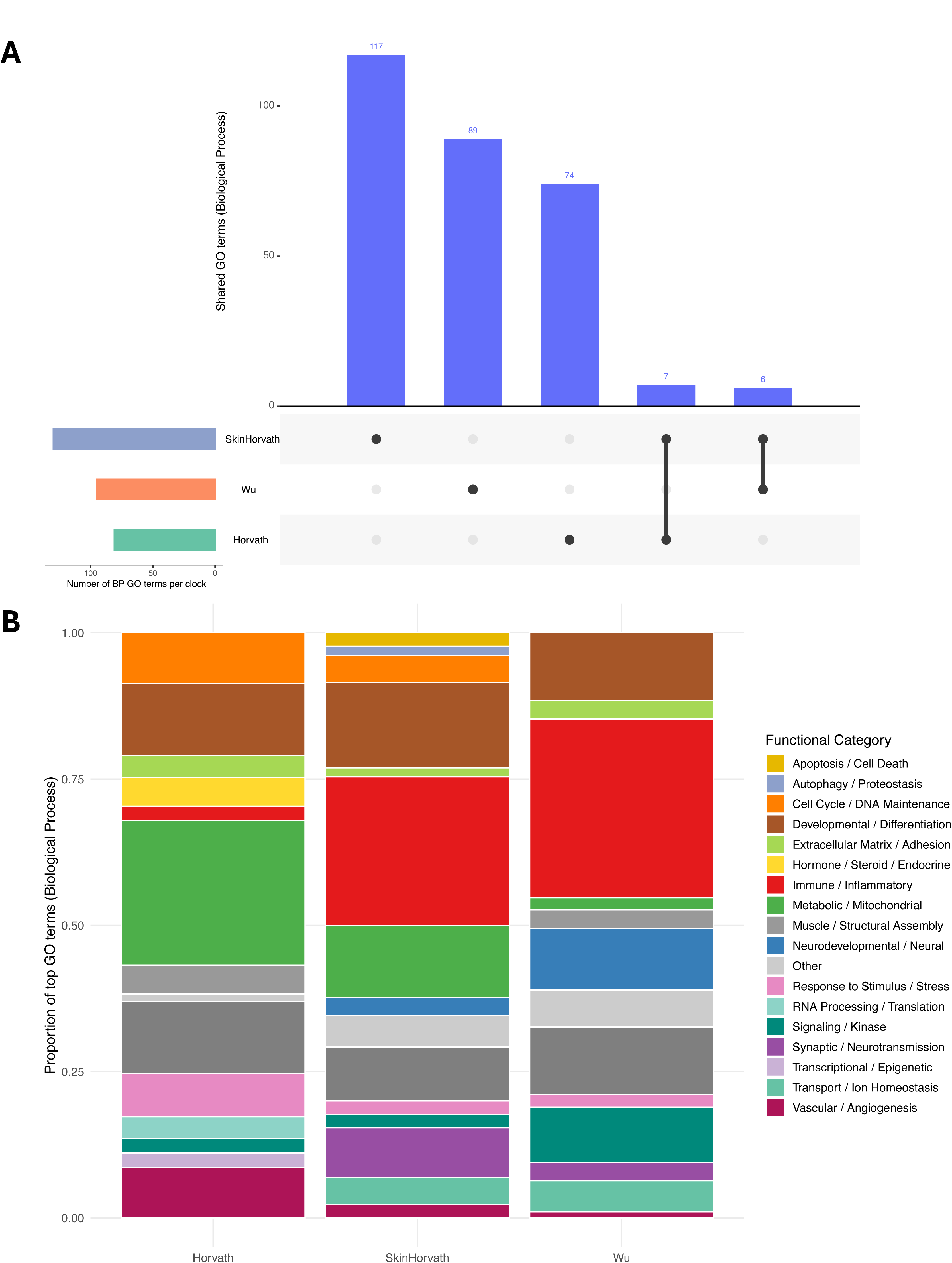
Gene ontology enrichment analysis of CpG sites compromising the Horvath, skinHorvath, and Wu epigenetic clocks. **A.** Upset plot showing the number enriched Biological Process (BP) GO terms unique to or shared across the Horvath, skinHorvath, and Wu clocks. Vertical bars, in blue, indicate the number of shared BP GO terms, and connected dots below indicate the corresponding clock combinations. Only BP GO terms with raw p < 0.05 and at least two annotated genes were included. **B.** Proportion of enriched GO terms grouped into broader functional categories for each clock. Functional categories were generated by grouping enriched GO terms based on shared biological themes.

With QGCOMP, the psychosocial factors mixture was associated with Horvath EAA, similar to WQS (Figure S5C). There was a 0.33-year increase in Horvath EAA per one quantile increase in the psychosocial factors (Ψ=0.33 years, 95% CI: 0.02, 0.63) (Figure S7). There was no association with skinHorvath EAA (Ψ=0.11 years, 95% CI: - 0.01,0.24) or Wu EAA (Ψ= 0.47 years, 95% CI: −0.04, 0.98), though the directions of effect were also positive.

No association was observed for any of the clocks in association with the pesticide metabolite mixture (Ψ [95% CI]: Wu: 0.13 (−0.06, 0.31); Horvath: −0.07 (−0.18, 0.06); skinHorvath: −0.003 [−0.05, 0.04] (Figure S5B).

### Sensitivity analysis

The associations between pesticide metabolite concentrations and longitudinal EAA additionally adjusting for cell-type proportions remained robust to the main analysis across all 3 clocks, with similar magnitudes of effect and precision (Table S1-S3). By contrast, associations for some psychosocial factors were modestly attenuated after cell-type adjustment. For example, for Wu EAA, the association with physical IPV decreased from β=0.18 years (95% CI: 0.06, 0.31) to β=0.13 years (95% CI: −0.01, 0.26). For Horvath EAA, the association with traumatic life events was attenuated from β=0.26 (95% CI: 0.12, 0.39) to β=0.19 (95% CI: 0.07, 0.32). Overall, effect sizes for psychosocial factors were reduced by approximately 15–30%, with loss of precision in some models. These findings suggest immune-cell composition may partially contribute to associations between psychosocial stressors and epigenetic aging.

The association between the joint mixture or pesticide mixture in association with longitudinal EAA remained consistent after dropping metabolites detected in less than 50% of samples (Figure S6). For the joint mixture, the effect estimates for Horvath EAA and skinHorvath EAA did not change, while the estimate for Wu EAA increased slightly (β= 0.43 years, 95% CI 0.19, 0.80). The top contributors remained the same. For the pesticide mixture, the effect estimates for Horvath decreased slightly (β= −0.03 years, 95% CI: −0.10, 0.02), while skinHorvath and Wu stayed the same. The top contributors in the main analysis for the Horvath clock and skinHorvath clocks were CINA6 and D24, respectively which were dropped from this analysis, but the rest of the weights remained consistent.

To assess whether the associations between prenatal exposures on EAA accumulated over time, we tested age x exposure interaction terms (Figure S7; Table S7-S9). Significant interactions were observed for food insecurity in association with skinHorvath clock, and for cortisol:cortisone, DMP, and D24 in association with the Wu clock (Figure S8). Marginal effects estimated at 12, 36, and 60 months consistently showed strengthening associations with increasing child age. Food insecurity was associated with a modest increase of 0.01 years of skinHorvath EAA per IQR at 12 months (β=0.01 (95% CI: −0.02, 0.05), which increased to 0.08 years at 60 months (β=0.08 (95% CI: 0.01, 0.14). Cortisol:Cortisone (β=-0.11 (95% CI: −0.25, 0.02), DMP β=0.01 (95% CI: - 0.15, 0.17), and D24 β=-0.21 (95% CI: −0.37, −0.05) pesticides were associated with decreased Wu EAA at 12 months, which increased at 60 months (Cortisol:Cortisone: β=0.19 (95% CI: −0.01, 0.39); DMP: β=0.26 (95% CI: 0.05, 0.48); D24: β=0.14 (95% CI: −0.08, 0.35).

We also tested whether associations between prenatal exposures and EAA differed by child sex (Figure S8; Table S10-S12). We observed significant sex-specific interactions for cortisol and cortisone in association with the Horvath clock and for DMDP in association with the skinHorvath clock (Figure S9). Higher cortisol and cortisone were associated with 0.19 years (95% CI: 0.08,0.30) and 0.14 years (95%CI: 0.05, 0.22) greater Horvath EAA in male children, but not in female children. Although the interaction was not statistically significant, higher cortisol:cortisone ratio was also associated with 0.11 years (95%CI: 0.005, 0.22) greater Horvath EAA in male children. Higher concentrations of DMDP were associated with increased sEAA among male children (β=0.07 (95% CI: −0.07, 0.21)) and decreased skinHorvath EAA among female children (β=-0.09 (95% CI: −0.22, 0.04).)

Associations were consistent across all five imputed datasets, with effect estimates showing similar magnitude and direction as the main analysis, indicating that findings were robust to the handling of missing data (Table S13-S15).

### Pathway enrichment analysis

After filtering Biological Process GO terms by a raw p-value < 0.05 and requiring at least two genes per term, we identified 117, 89, and 74 unique GO terms, for the skinHorvath, Wu, and Horvath clocks, respectively (Figure 4a; Table S16-S18). Overlap across clocks was limited, with seven terms shared between skinHorvath and Horvath, six between skinHorvath and Wu, and none between Wu and Horvath (Figure 4a; Table S19). After grouping the terms into broader functional categories, we identified distinct underlying architectures for each clock (Figure 4b; Table S20). For the Horvath clock, the most represented categories were Metabolic/Mitochondrial processes (24.7%), Developmental/Differentiation (12.3%), and Protein Homeostasis/Modification (12.3%). For the skinHorvath clock, Immune/Inflammatory pathways comprised the largest proportion (25.4%), followed by Developmental/Differentiation (11.6%) and Metabolic/Mitochondrial (12.3%). For the Wu clock, the most represented categories were Immune/Inflammatory pathways (30.5%), Developmental/Differentiation (11.6%), and Protein Homeostasis/Modification (11.6%). The Wu clock additionally showed a higher proportion of Neurodevelopmental pathways (10.5%) compared with the Horvath (0%) and skinHorvath (3.1%) clocks. Together, these findings may partly explain the distinct associations between environmental factors and these three epigenetic clocks, pointing to potential biological pathways that link early-life environments to aging and development.

## Discussion

In this prospective, longitudinal South African birth cohort, we found evidence that joint prenatal exposure to pesticides, as measured using urinary metabolites, and psychosocial factors was associated with increased EAA across early childhood. Using complementary mixture modeling approaches, including WQS regression and QG-COMP, we observed that the joint exposures were associated with increased EAA over time. Across analyses, exposure to psychosocial factors alone or in combination with pesticides was more strongly associated with EAA than pesticide exposure alone. Taken together, these findings indicate that co-occurring prenatal psychosocial factors and pesticide exposures jointly contribute to variation in EAA during early childhood. Pathway enrichment analyses of CpGs present in different epigenetic clocks revealed clock-specific biological architectures, with the Wu clock enriched for neurodevelopmental and immune pathways, the skinHorvath clock for immune and cell differentiation, and the Horvath clock for metabolic and cell differentiation pathways. The clock-specific biological architecture may partly explain the distinct associations with environmental factors.

Longitudinal studies linking early-life environmental exposures to EAA in children remain limited, but available studies suggest that epigenetic aging trajectories may be particularly sensitive to prenatal exposures. In one of the earliest studies, Simpkin et al. reported that epigenetic age acceleration varied across childhood and was associated with early-life factors, with some differences resolving during childhood^54^. Consistent with these findings, we observed stronger associations between prenatal exposures and EAA in the first three years of life, with attenuation by five years, suggesting that the impact of exposures on epigenetic aging signals in relation to prenatal exposures may be most important during fetal life and early development.

A longitudinal study in the Project Viva cohort examined prenatal maternal social experiences in association with offspring EAA in the first 7 years of life and found that increased maternal experiences of discrimination were associated with decreased offspring EAA in early to mid-childhood, but not at birth^55^. One longitudinal study from the CHAMACOS cohort assessed associations between prenatal chemical and non-chemical stressors and EAA in childhood^56^. They found that children whose mothers engaged in agricultural fieldwork exhibited increased Horvath and skinHorvath EAA during adolescence, independent of pesticide exposure, suggesting that agricultural work may serve as a proxy for psychosocial stressors^56^. This aligns with our findings in which psychosocial factors, either alone or jointly with urinary pesticide metabolite levels, were more strongly associated with EAA than any individual pesticide metabolite. Nevertheless, associations between prenatal exposures and epigenetic aging are likely not uniform across developmental stages, exposure domains, or epigenetic clocks, and may manifest differently during sensitive periods such as early childhood or puberty^57,58^.

Among the pesticide metabolites we evaluated, PBA and TDCCA were the largest contributors to Wu EAA across both single-exposure and mixture models. PBA and TDCCA are complementary urinary metabolites of the same parent pyrethroid insecticides, primarily permethrin (type I) and cypermethrin (type II), and are extensively used in agricultural and household settings for pest control^59^. In the Western Cape, which is a major fruit and wine-producing area, permethrin and cypermethrin were detected at the highest concentrations among pesticides in air and dust samples in neighborhoods near Western Cape farms, showing strong evidence of pesticide drift from neighboring vineyards^60^. While pyrethroids are established neurotoxicants and endocrine disruptors, the epigenetic mechanisms underlying the association remain unclear^61^. Future studies should investigate the specific biological pathways through which prenatal pyrethroid exposure may influence epigenetic aging in childhood.

Our results underscore the importance of considering the joint effects of both chemical and non-chemical stressors when evaluating early-life influences on epigenetic aging. In stratified analyses, the psychosocial factors mixture exhibited a larger and more precise effect estimate in comparison to the pesticide mixture alone, suggesting that psychosocial stressors may account for a substantial proportion of the observed association with EAA. We note that with WQS, the joint exposure mixture was also significantly associated with EAA and exhibited a larger effect estimate than either psychosocial factors or pesticides alone, demonstrating the importance of assessing cumulative exposure burden rather than isolated exposures.

These findings are consistent with emerging evidence suggesting that psychosocial stressors may amplify susceptibility to environmental toxicants. Animal and *in vitro* studies have suggested that stress can exacerbate the neurotoxic effects of pesticides through glucocorticoid-mediated mechanisms, thereby sensitizing developing neural circuits to subsequent disruption by pesticides^62–64^. Few epidemiologic studies have identified associations between joint exposures to environmental and psychosocial stressors and child brain health outcomes^65–67^. However, to our knowledge, no study has evaluated joint exposures in relation to epigenetic aging, either at one time point or longitudinally.

Across single exposures, mixtures, and interaction analyses, the glucocorticoid stress biomarkers (cortisol, cortisone, and cortisol:cortisone ratio), were consistent contributors to EAA, suggesting that prenatal HPA axis dysregulation may be an important pathway linking prenatal exposures to childhood EAA, consistent with prior evidence that maternal prenatal stress is associated with differential DNA methylation in infants^68^. The cortisol:cortisone ratio acts as a biomarker of 11β -HSD2 enzyme activity, which converts active cortisol to inactive cortisone, and has been found to protect the developing fetus from excess maternal glucocorticoid exposure^69^. A higher ratio may reflect greater fetal glucocorticoid levels and maternal psychosocial stress burden. Prenatal psychosocial stress has also been shown to epigenetically downregulate placental 11β -HSD2 expression^70^. In this study, cortisol:cortisone ratio showed a strengthening association with Wu EAA over the first five years of life. Sex-stratified analyses showed associations between cortisol, cortisone, and Horvath EAA among male children. Prior studies have established that male fetuses may be more sensitive to exposure to high maternal cortisol, including differential methylation of 11B-HSD2, whereas female fetuses may exhibit adaptive mechanisms^71,72^. Future studies should examine whether glucocorticoid stress biomarkers mediate or moderate the association between prenatal psychosocial stressors, environmental exposures, and EAA to assess whether glucocorticoid regulation is a potential biological pathway linking the prenatal environment to early-life epigenetic aging.

As in prior studies, we observed heterogeneity in associations across epigenetic clocks. In this study, pesticides and psychosocial exposures were most consistently associated with Wu EAA, whereas associations with Horvath and skinHorvath EAA were more limited and primarily observed for psychosocial factors. While heterogeneity may in part be explained by differences in prediction accuracy and training populations, the differences in the underlying biological architecture captured by each clock has not been considered. In our pathway enrichment analysis, the Wu clock was characterized by a larger proportion of CpG sites mapping to neurodevelopmental and immune-related biological processes compared with Horvath and skinHorvath clocks, which were more heavily enriched for metabolic, mitochondrial, and a smaller proportion of immune-related pathways. The limited overlap in enriched GO terms across clocks further demonstrates that epigenetic clocks are not a unitary construct, and that clock-specific biological architecture may explain the distinct associations between early-life exposures and different epigenetic clocks. Given its enrichment for neurodevelopmental processes, the Wu clock may be particularly sensitive to early-life environmental exposures occurring during a critical period for brain development. Future research should investigate whether clock-specific biological architecture can inform the development of targeted pediatric epigenetic clocks, and whether EAA in early childhood differentially predicts later neurodevelopmental and health outcomes across clock types.

Despite its strengths, our study has several limitations. The pesticides evaluated in this study have short biological half-lives thus a single urine sample collected during pregnancy may not accurately capture average exposure across pregnancy. Given that this cohort was likely occupationally exposed, we expect that one timepoint will likely capture average exposure as has been demonstrated in other occupational (farmworker) birth cohorts^73^. In addition, many of the metabolites evaluated (e.g., PBA, TDCCA, DEP, DMP) can be preformed in the environment such that co-exposure may occur to the active pesticide and inactive metabolite. However, in occupational settings, environmental concentrations of the intact pesticides are far greater than the preformed metabolite thus reducing the likelihood of overestimation of exposure^73^. Additionally, some of the pesticide metabolites had low detection levels. Although we imputed levels below the detection limit using left-censoring, low detection may reduce power to detect associations for some pesticides and may bias our results towards the null. Finally, unmeasured childhood exposures may contribute to variability in epigenetic aging during childhood, which limits our ability to disentangle prenatal from early-life effects.

This study is among the first to investigate epigenetic age acceleration measured at multiple early-life timepoints in relation to prenatal mixtures of chemical and non-chemical exposures, using several advanced environmental mixture methods to characterize these complex relationships. By leveraging rich longitudinal epigenetic data in an underserved LMIC population with high exposure burdens, our findings provide novel insights into how environmental and psychosocial risk factors jointly influence biological aging processes during early childhood—a period of rapid development and heightened susceptibility. To our knowledge, this is also among the first studies to characterize the biological processes underlying pediatric epigenetic clocks through pathway enrichment analysis, revealing clock-specific architectures that may help explain heterogeneity in associations across clocks. Understanding how cumulative prenatal exposures shape epigenetic aging trajectories may help identify vulnerable subgroups and inform early-life strategies aimed at reducing health disparities.

## Data Availability

All data produced in the present study are available upon reasonable request to the authors.

## Funding and Acknowledgements

The authors would like to thank the study and clinical staff at Paarl Hospital, Mbekweni and TC Newman clinics, as well as the CEO of Paarl Hospital, and the Western Cape Health Department for their support of the study. The authors also thank the families and children who participated in this study.

The Drakenstein Child Health Study was funded by the Bill & Melinda Gates Foundation (OPP 1017641 and OPP1017579), the Wellcome Trust (221372/Z/20/Z)), with additional support for this work from the South African Medical Research Council and the National Research Foundation of South Africa.

SA was supported by the National Institute of Environmental Health Sciences (T32ES012870) and (F31ES037540). AAL is supported by an MQ Fellows Award (MQF22\9) and awards from the National Institutes of Health (R21MH132947; R21AA030640). DBB was funded in part by P30ES019776.

The National Institutes of Health/National Institute of Environmental Health Sciences has awarded a grant to the Mount Sinai HHEAR Targeted Analysis Laboratory (U2CES026561, PI: Robert O. Wright) for the laboratory analysis of urine creatinine, pesticide metabolites, and biomarkers of psychosocial stress.

The epigenetic data described in this report from the DCHS were supported by the National Institute of Mental Health of the National Institutes of Health (R01MH113930, PI: ECD).

The NIH had no further role in study design; in the collection, analysis and interpretation of data; in the writing of the report; and in the decision to submit the paper for publication. The content is solely the responsibility of the authors and does not necessarily represent the official views of the National Institutes of Health.

