## Supplemental Figures for "The joint effects of exposure to prenatal pesticides and psychosocial factors on epigenetic age acceleration in the first 5 years of life in a South African birth cohort"

### Supplementary Figures

Sarina Abrishamcar^1^, Stephanie M. Eick^1,2^, Todd Everson^1,2^, Shakira F. Suglia^1^, M. Daniele Fallin^1^, Robert O. Wright^3^, Syam S. Andra^3^, Jasmin Chovatiya^3^, Ravikumar Jagani^3^, Dana Boyd Barr^2^, Alexandre A. Lussier^4,5,6^, Erin C. Dunn^7^, Julie L. MacIsaac^8,9^, Kristy Dever^8,9^, Michael S. Kobor^8,9^, Nadia Hoffman^10,11^, Nastassja Koen^10,12^, Heather J. Zar^13^, Dan J. Stein^†10,11^, Anke Hüls*^1,2,14^

^1^Department of Epidemiology, Rollins School of Public Health, Emory University, Atlanta, GA, USA

^2^Gangarosa Department of Environmental Health, Rollins School of Public Health, Emory University, Atlanta, GA, USA

^3^Department of Environmental Medicine, Icahn School of Medicine at Mount Sinai, New York, NY 10029, USA.

^4^Center for Genomic Medicine, Massachusetts General Hospital, Boston, MA, USA

^5^Department of Psychiatry, Harvard Medical School, Boston, MA, USA

^6^Division of Depression & Anxiety Disorders, McLean Hospital, Belmont, MA, USA

^7^Department of Sociology, College of Liberal Arts, Purdue University, West Lafayette, IN, USA

^8^Edwin S. H. Leong Centre for Healthy Aging, Faculty of Medicine, University of British Columbia, Vancouver, BC, Canada

^9^British Columbia Children's Hospital Research Institute and Department of Medical Genetics, University of British Columbia , Vancouver, BC, Canada

^10^Neuroscience Institute, University of Cape Town, Cape Town, South Africa

^11^South African Medical Research Council (SAMRC) Unit on Risk and Resilience in Mental Disorders, University of Cape Town, Cape Town, South Africa

^12^Department of Psychiatry and Mental Health, University of Cape Town, Cape Town, South Africa

^13^Department of Paediatrics and Child Health, SA-MRC unit on Child & Adolescent Health, Red Cross War Memorial Children’s Hospital, University of Cape Town, Cape Town, South Africa

^14^Department of Biostatistics and Bioinformatics, Rollins School of Public Health, Emory University, Atlanta, GA, USA

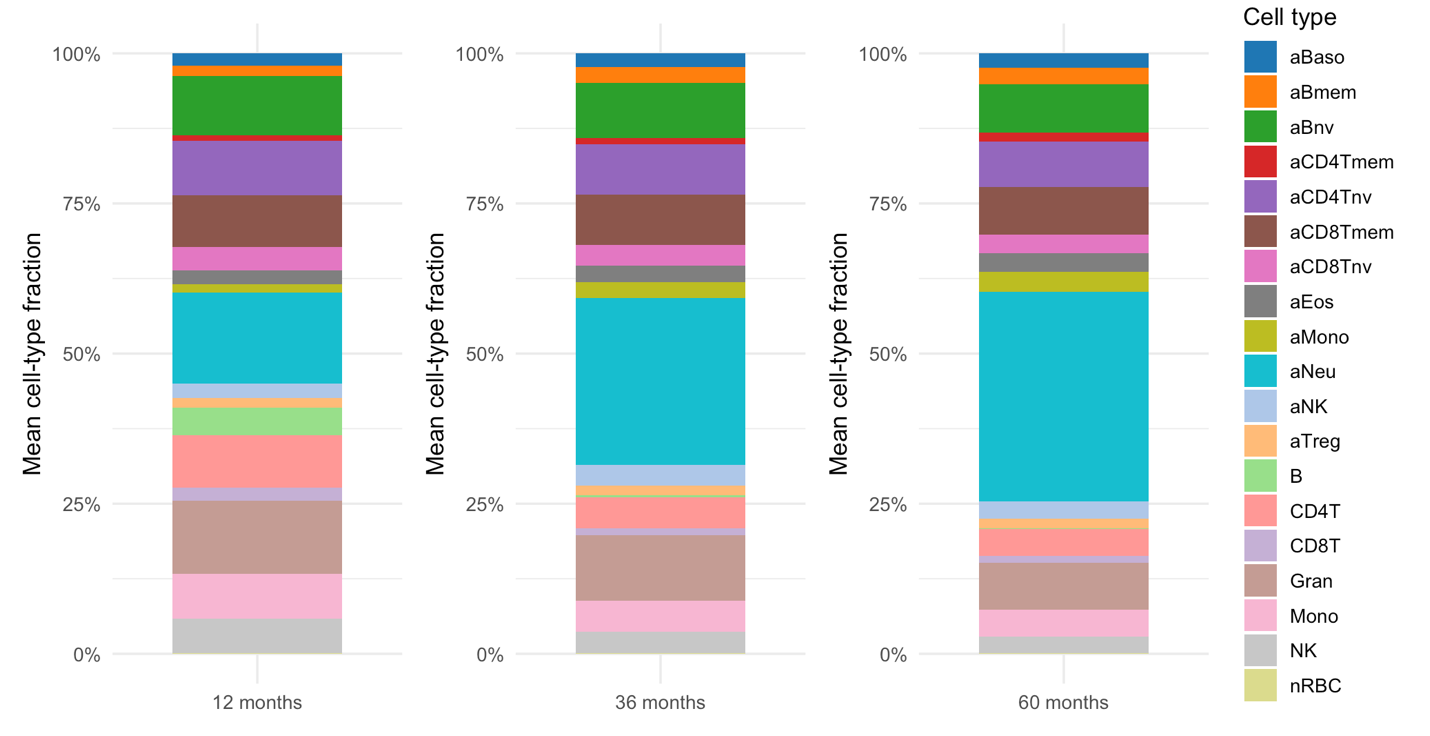

**Figure S1. Estimated blood immune cell-type proportions across early childhood.** Estimated immune cell-type proportions at 12, 36, and 60 months derived using the UniLife reference panel implemented in EpiDISH. UniLife predicts 19 immune cell-types including seven cord-blood subtypes (B cells, NK cells, granulocytes, monocytes, nRBCs, CD4T, and CD8T cells) and 12 adult immune cell-types (naïve and mature B cells, naïve and mature CD4T cells, naïve and mature CD8T cells, T-regulatory cells, NK cells, neutrophiles, monocytes, eosinophils, basophils). Proportions reflect predicted fractions of cord-blood and adult immune cell subtypes. Cell types with prefix “a” are adult cell types.

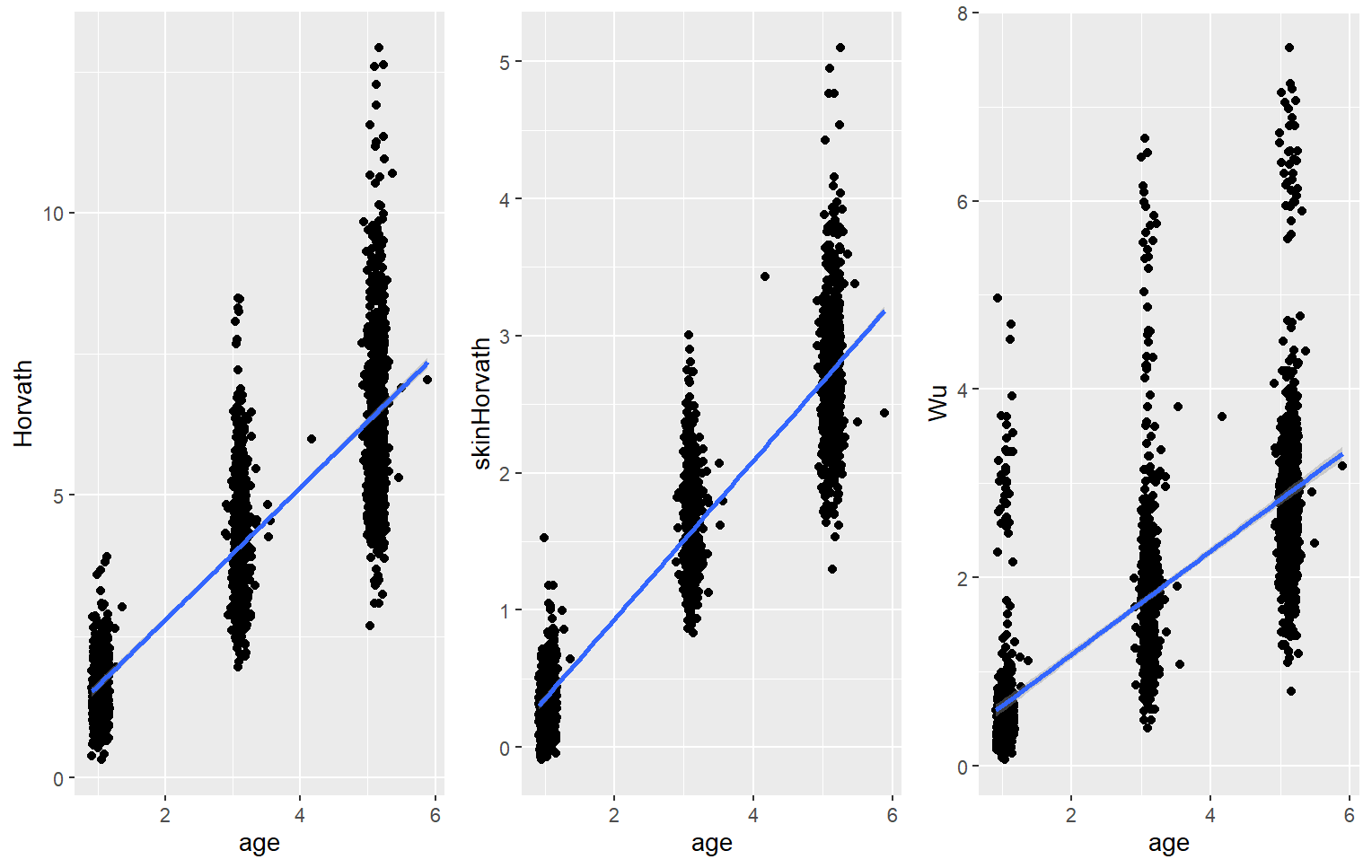

R^2^=0.86
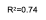

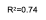

R^2^=0.51
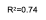

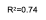

R^2^=0.74

**Figure S2: Correlation between epigenetic age and chronological age.** Scatterplots showing the correlation between epigenetic age and chronological age at 12, 36, and 60 months for the Horvath, skinHorvath, and Wu clocks. Lines represent fitted linear regression models. R² values are displayed for each clock at each timepoint

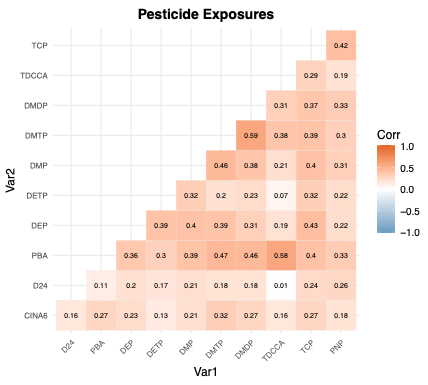

**Figure S3. Correlation matrix of prenatal urinary pesticide metabolites.** Pearson correlation matrix of prenatal urinary pesticide metabolites measured during the second trimester of pregnancy. Color intensity corresponds to the strength and direction of correlation coefficients.

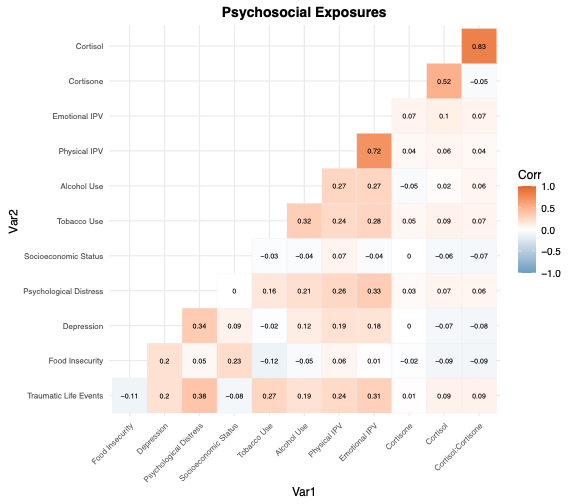

**Figure S4. Correlation matrix of prenatal psychosocial factors.** Pearson correlation matrix of prenatal psychosocial factors assessed during the second trimester of pregnancy. Color intensity corresponds to the strength and direction of correlation coefficients.

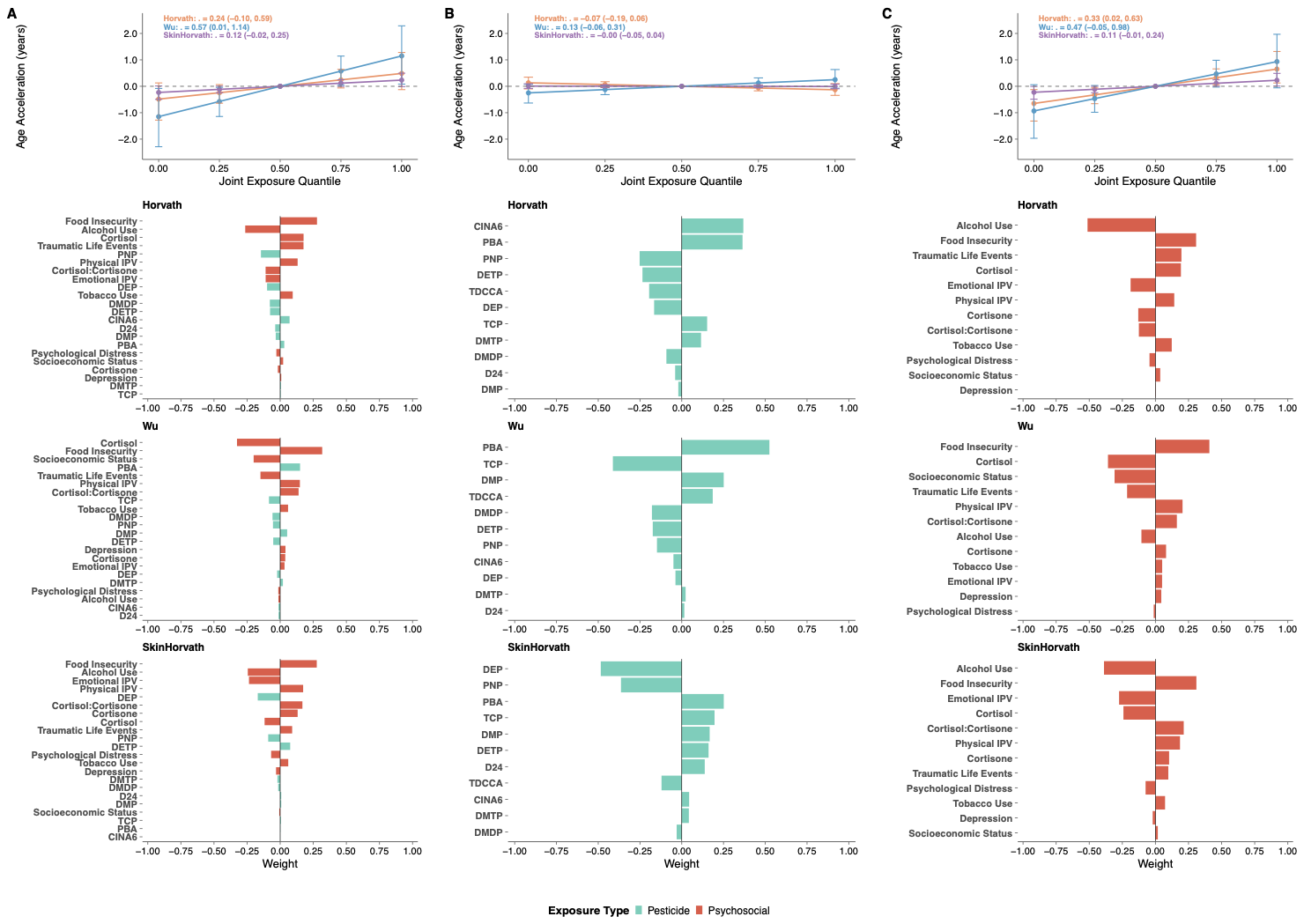

**Figure S5. Quantile g-computation (QGCOMP) mixture associations between the joint prenatal exposure mixture and epigenetic age acceleration over time.** Longitudinal associations between the combined prenatal pesticide and psychosocial exposure mixture and epigenetic age acceleration (EAA) across 12, 36, and 60 months. **Column A.** Association between the joint prenatal exposure mixture and EAA across 12, 36, and 60 months. **Column B.** Longitudinal associations between pesticide metabolite mixtures alone and EAA. **Column C**. Longitudinal associations between psychosocial factor mixtures alone and EAA.

Results are shown from the longitudinal generalized estimating equations QG-COMP model. For each column, the top plot displays the estimated cumulative effect (Ψ) representing the change in EAA (years) per one quintile increase in all exposures simultaneously, along with 95% confidence intervals and model-based confidence bands. Lower plots display the mixture weights, separated into positive and negative weights, indicating the relative contribution and direction of each exposure to the overall mixture effect. Weights sum to the total positive and negative components within each model. All models were adjusted for child age, child sex, race, maternal age at enrollment, and maternal HIV status. Models including pesticide metabolites or psychosocial biomarkers were additionally adjusted for urinary creatinine. Pesticide metabolites are shown in blue and psychosocial factors are shown in salmon.

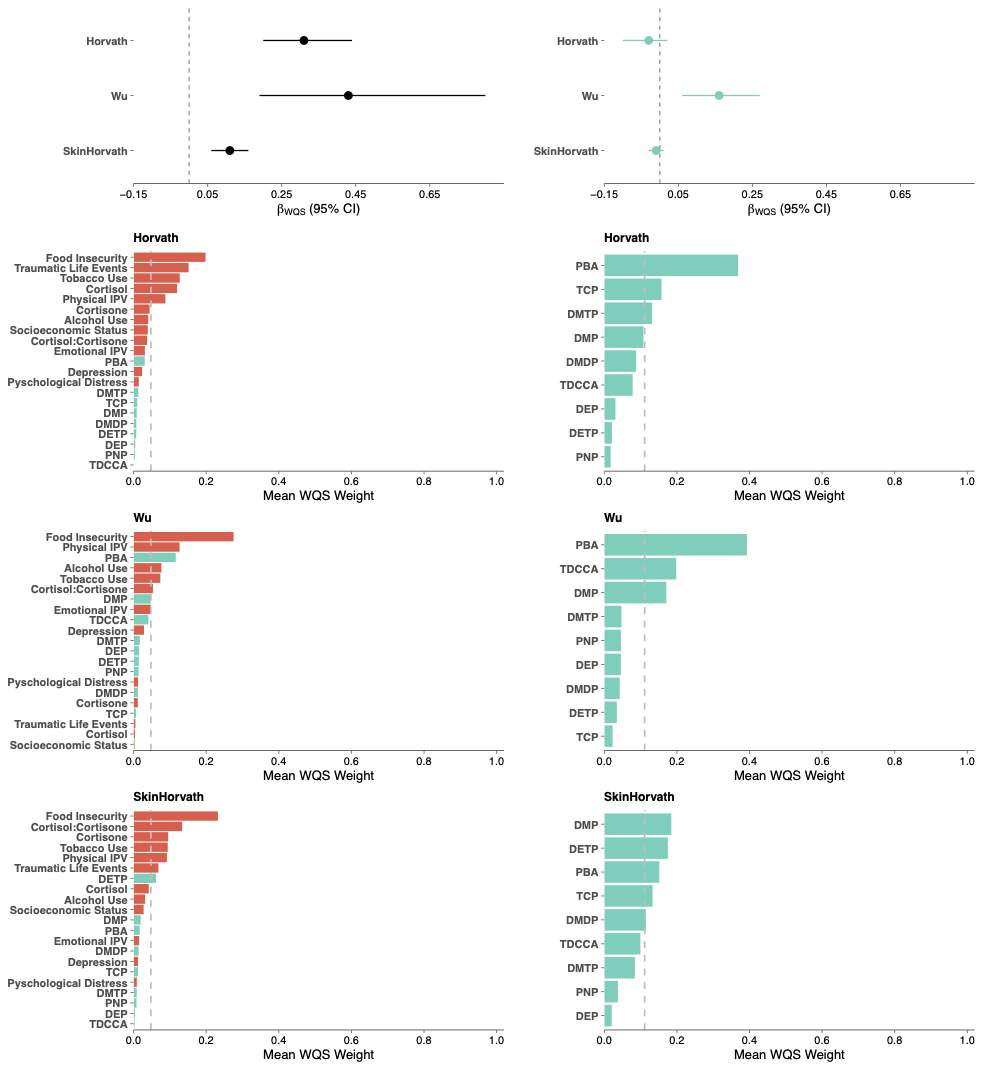

**Figure S6: Weighted quantile sum (WQS) mixture associations between prenatal pesticide metabolites and psychosocial factors and epigenetic age acceleration (Horvath, Wu, and Skin and Blood Horvath) during follow-up, after dropping pesticides that were detected in less than 50% of samples (CINA6 and D24).** **Column A.** Association between the joint prenatal exposure mixture and EAA across 12, 36, and 60 months. **Column B.** Longitudinal associations between pesticide metabolite mixtures alone and EAA.

Results are shown from the longitudinal mixed-effects WQS regression with repeated holdout validation. Beta estimates represent the change in EAA (years) per one quintile increase in the exposure. Point estimates denote pooled effect estimates (median across 100 repeated holdouts) and the corresponding 95% confidence intervals (2.5th–97.5th percentiles). For each column, lower plots display the corresponding WQS mixture weights indicating the relative contribution of each exposure to the overall mixture effect. Weights sum to one within each model. Pesticide metabolites are shown in blue and psychosocial factors are shown in salmon. All models were adjusted for child age, child sex, race, maternal age at enrollment, maternal HIV status, and creatinine.

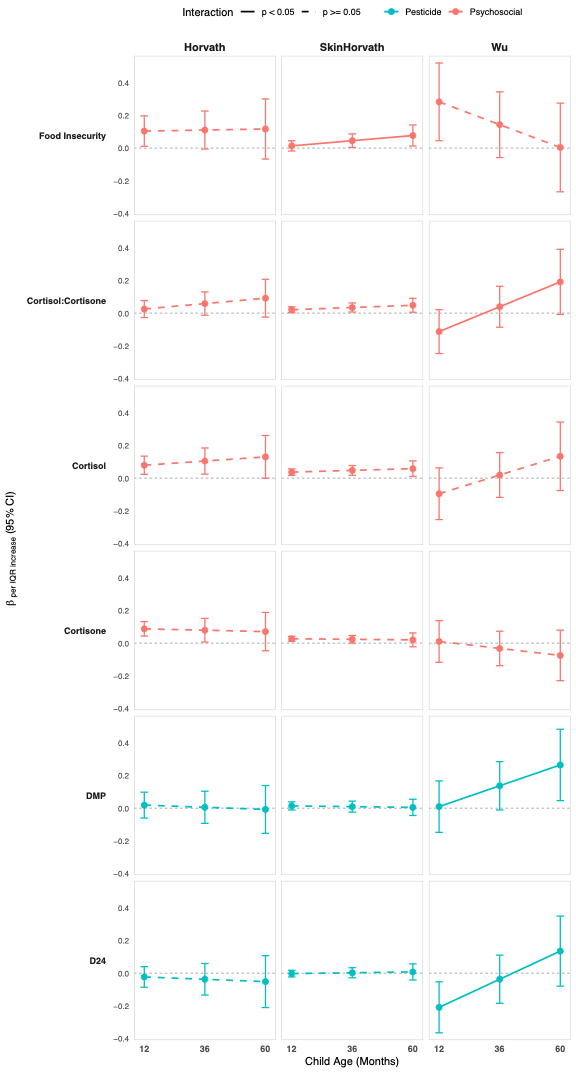

**Figure S7:** **Age-stratified marginal effects of prenatal exposures on EAA during follow-up.** Marginal effects estimated at 12, 36, and 60 months from generalized estimating equations (GEE) models including an exposure x age interaction term. Exposures are shown if they had a statistically significant interaction with child age (p < 0.05) for at least one epigenetic clock. Beta coefficients represent the change in EAA per one interquartile range (IQR) increase in log-transformed exposure at each age. Solid lines indicate statistically significant associations (p < 0.05) and dashed lines indicate non-significant associations (p ≥ 0.05). Psychosocial factors are shown in salmon and pesticide metabolites are shown in blue. All models were adjusted for child sex, race, maternal age at enrollment ,and maternal HIV status. Models including pesticide metabolites or psychosocial biomarkers were additionally adjusted for urinary creatinine.

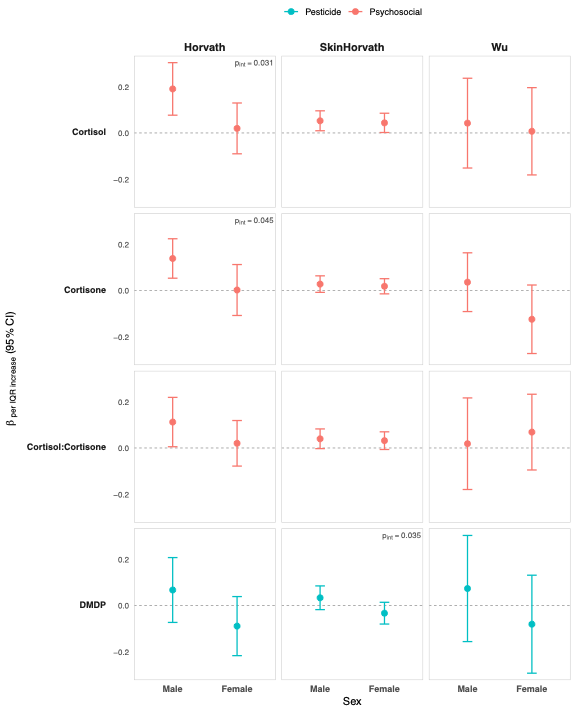

**Figure S8:** **Age-stratified marginal effects of prenatal exposures on EAA during follow-up.** Marginal effects estimated separately for male and female children from generalized estimating equations (GEE) models including an exposure × sex interaction term. Exposures are shown if they had a statistically significant interaction with child sex (p < 0.05) for at least one epigenetic clock. Beta coefficients represent the change in epigenetic age acceleration (EAA, in years) per one interquartile range (IQR) increase in log-transformed exposure. Interaction p-values are displayed only where the exposure × sex interaction was statistically significant (p < 0.05). Psychosocial factors are shown in salmon and pesticide metabolites are shown in blue. All models were adjusted for child age, race, maternal age at enrollment, maternal HIV status. Models including pesticide metabolites or psychosocial biomarkers were additionally adjusted for urinary creatinine.
